# ATTCT and ATTCC repeat expansions in the *ATXN10* gene affect disease penetrance of spinocerebellar ataxia type 10

**DOI:** 10.1101/2022.05.12.22274972

**Authors:** C. Alejandra Morato Torres, Faria Zafar, Yu-Chih Tsai, Jocelyn Palafox Vazquez, Michael D. Gallagher, Ian McLaughlin, Karl Hong, Jill Lai, Joyce Lee, Amanda Chirino-Perez, Angel Omar Romero-Molina, Francisco Torres, Juan Fernandez-Ruiz, Tetsuo Ashizawa, Janet Ziegle, Francisco Javier Jiménez Gil, Birgitt Schüle

**Author notes:** Corresponding author: Birgitt Schüle. These authors contributed equally to this work (C. Alejandra Morato Torres, Faria Zafar).

## Abstract

Spinocerebellar ataxia type 10 (SCA10) is an autosomal-dominant disorder caused by an expanded pentanucleotide repeat in the *ATXN10* gene. This repeat expansion, when fully penetrant, has a size of 850 to 4500 repeats. It has been shown that the repeat composition can be a modifier of disease, e.g., seizures.

Here, we describe a Hispanic kindred in which we identified both pure (ATTCT)_n_ expansions and mixed (ATTCT)_n_-(ATTCC)_n_ in the same family. We used No-Amp targeted sequencing and optical genome mapping to decipher the composition of these repeat expansions. We found a considerable degree of mosaicism in the repeat expansion. This mosaicism was confirmed in skin fibroblasts from *ATXN10* carriers with RNAScope in situ hybridization. All affected family members with the mixed *ATXN10* repeat expansion showed typical clinical signs of spinocerebellar ataxia and epilepsy. In contrast, individuals with the pure *ATXN10* expansion present with Parkinson’s disease or are unaffected even more than 20 years older than the average age at onset for SCA10.

Our findings suggest that the pure (ATTCT)_n_ expansion is non-pathogenic while repeat interruptions, e.g., (ATTCC)_n_, are necessary to cause SCA10. This mechanism has been recently described for several other repeat expansions, including SCA31 (*BEAN1*), SCA37 (*DAB1)*, and three loci for benign adult familial myoclonic epilepsy BAFME (SAMD12, TNRC6A, RAPGEF2). Therefore, long-read sequencing and optical genome mapping of the entire genomic structure of repeat expansions is critical for clinical practice, and genetic counseling as variations in the repeat can affect disease penetrance, symptoms, and disease trajectory.

## Introduction

Complex pentanucleotide repeats within intron 9 of the *ATXN10* gene cause spinocerebellar ataxia type 10 (SCA10), a rare autosomal-dominant disease that is most prevalent in Middle and South America^1^. These repeats can comprise several thousand when clinically fully penetrant ^2; 3^. The normal ATTCT repeat allele size is 10–32 repeats, whereas intermediate alleles with reduced penetrance contain 280–850 ATTCT repeats. Complete penetrance of SCA10 is typically seen between 850 and 4500 ATTCT repeats ^2; 3^.

Clinically, SCA10 presents with a symptom complex of spinocerebellar ataxia, gait ataxia, dysarthria, nystagmus, epilepsy, cognitive decline, and non-motor symptoms^4^. These symptoms affect quality of life measures, particularly “physical functioning” and “physical role”.^5^ In addition, non-motor symptoms include increased fatigue (32% of SCA10 versus 3.6% control group (p=0.005)^6^ and changes in rapid eye movement (REM) sleep pattern (more extended REM periods and more REM arousals) and a higher respiratory disturbance index (RDI)^7^. Assessment of changes in the sense of smell or cancer risk shows no significant differences compared to controls^8; 9^. High-resolution magnetic resonance imaging (MRI) in 18 SCA10 patients revealed white matter atrophy starting in the posterior/ floculonodular cerebellum and grey matter degeneration in the cerebellum and brainstem thalamus. The putamen was more pronounced in patients with severe disease status. Additionally, SCA10 patients with epilepsy had lower grey matter intensity in the thalamus and a loss of integrity of the white matter in the lobule VI of the cerebellum.^10^

Diagnostically, the genetic sequencing analysis of *ATXN10* expansion length has been challenging due to its repetitive nature. Furthermore, Southern blotting has inherent problems with accurate sizing and considerable variability between clinical diagnostic laboratories. PCR for repetitive regions shows reduced amplification efficiency, allele bias, allele dropout^11^, and replication slippage effects^12^. Therefore, the somatic variability of repeat elements is challenging to assess because of replication errors.

Besides the repeat length, the *ATXN10* pentanucleotide repeat can have variable genomic structure configurations: pure (ATTCT)n repeats and mixed (ATTCT)n-(ATTCC)n or (ATTCT)n-(ATCCT)n-(ATCCC)n with additional interruptions of hepta- or octanucleotide repeats^3^. However, current clinical genetic diagnostics cannot determine the *ATXN10* pentanucleotide repeat composition. More recent developments of repeat-prime PCR with the high-resolution pulse-field capillary electrophoresis analysis^13^ and long-range sequencing^3^ in the research setting allow for refinement of *ATXN10* repeat composition.

It is very likely that the genomic composition of the *ATXN10* repeat expansion leads to different clinical phenotypes and pathophysiological changes in patients and may influence the pathogenicity and penetrance of the expanded repeat allele. E.g., epileptic seizures in SCA10 can present as complex partial with occasional secondary generalization greatly influencing morbidity and mortality of the disease. The AT<u>C</u>CT interruption of the ATTCT repeat expansion shows a higher risk (6.3 fold) of developing seizures^14^.

Here, we performed No-Amp targeted sequencing (Pacific Biosciences) and optical genome mapping (Bionano Genomics) in three generations of a large Hispanic family with SCA10 to assess repeat size and repeat composition in affected and unaffected relatives. We characterized the somatic variability of the repeat composition in skin fibroblasts from *ATXN10* carriers. We could show that the variable expansions translated into variably size and number of RNA foci detected by RNAScope *in situ* hybridization.

## Subjects and Methods

### Ethics statement

This study was approved by the ethics committee of Stanford University School of Medicine (Stanford IRB-48895). All participants provided written informed consent.

### Clinical assessment

We previously reported on this family and have expanded the recruitment of family members to include relatives of the third-generation^15^. Cases histories for family members that carry repeat expansions but have not been previously reported are described in the **Supplemental Data**.

### DNA isolation

DNA from saliva extracted with Oragene-500 kit according to manufacturer’s instructions. For optical genome mapping, we isolated ultra-high molecular weight (UHMW) genomic DNA from blood using the Bionano Prep SP Blood & Cell Culture DNA Isolation Kit (Bionano, Cat. No. 80030).

### Short tandem repeat (STR) fragment analysis for identity testing

For identity testing, we used the AmpFlSTR™ Identifiler™ Plus PCR Amplification Kit (Thermo Fisher, Cat. No. 4427368) according to the manufacturer’s instructions. The kit uses a five-dye fluorescent system, and the inclusion of non-nucleotide linkers allows for simultaneous amplification and efficient separation of 15 short tandem repeat loci. Electropherograms of the fragments were analyzed using NCBI’s OSIRIS 2.13.1 program (**Supplemental Table 1**).

### No-Amp targeted sequencing

DNA samples were further purified with AMPure PB beads (PacBio, Cat. No. 100-265-900) before processing, and quality control (QC) on DNA samples was performed using Agilent Femto pulse analysis. Using a modified PacBio No-Amp targeted sequencing method protocol, we started with five µg input DNA for each sample and sequenced with 20 hrs of PacBio Sequel System/Sequencing run. More specifically, non-sequenceable SMRT (Single Molecule, Real-Time) bell libraries were generated from native genomic DNA digested with the restriction enzyme, EcoRI-HF, and a short hairpin adapter with barcode sequences. These short hairpin adapters do not contain sequencing primer complementary sequence to prevent sequencing reaction initiation. We then introduced double-strand breaks to the SMRTbell templates that contained our region of interest using Cas9 and a guide RNA (*ATXN10*-5’-AUACAAAGGAUCAGAAUCCC-3’) designed to be complementary to a sequence adjacent to the region of interest ^16^. The SMRT bell library hairpin adapter was subsequently ligated to the digested templates to form asymmetric SMRT bell libraries that allow sequencing primer to anneal with the second hairpin adapter introduced after Cas9 enzyme digestion. The library was digested with an enzyme cocktail containing exonuclease III, exonuclease VII, and four restriction enzymes (BssSαI, BlnI, SpeI, KpnI) to remove a large portion of non-target DNA fragments from the final library. The selected restriction enzymes are not predicted to cut inside the DNA fragments with the *ATXN10* repeat expansion region. The final sequencing libraries were treated with Trypsin and purified with AMPure PB beads before primer annealing, polymerase binding, and sequencing.

Sequencing data were first processed with PacBio SMRT analysis tool version 5.0 to identify hairpin adapters, demultiplex reads from different samples, and generate circular consensus reads (CCS reads) with predicted read quality above Q20 Phred quality score. CCS reads from each sample were mapped against human reference sequence hg19, and ATXN10 repeat sequences from on-target sequencing reads were extracted from reads mapped through reference coordinates chr22:46191235-46191304. Repeat sequence lengths distribution and waterfall plots depicting repeat sequence variations (**Figure 2, Supplemental Figure 1**) were generated using either custom software scripts or repeat analysis tools available on the GitHub website (https://github.com/PacificBiosciences/apps-scripts/tree/master/RepeatAnalysisTools) following the recommended data analysis workflow on PacBio website (https://www.pacb.com/wp-content/uploads/Analysis-Procedure-No-Amp-Data-Preparation-and-Repeat-Analysis.pdf).

### Optical genome mapping and analysis with the Saphyr system

Briefly, 750ng ultra-high molecular weight (>100kb) genomic DNA was labeled with DLE-1 enzyme (Bionano Genomics) and DNA backbone counterstained according to Bionano Prep Direct Label and Stain protocol (Document Number: 30206 Document Revision: F) before loading on to the Bionano Chip. Loading of the chip and running of the Bionano Genomics Saphyr System were performed according to the Saphyr System User Guide (https://bionanogenomics.com/support-page/saphyr-system/). We used the Bionano de-novo assembly and variant annotation pipelines to generate whole-genome assembly contigs which span whole chromosome arms and identify variants down to 500bp. For single variant (SV) detection, the consensus genome maps were aligned to reference (hg38). After running the de novo assembly pipeline using Solve v1.7 and visualizing with Access v3.7, we identified somatic rearrangements by selecting novel SVs to the Bionano control sample database (zero population frequency [out of 204 samples], thus potentially somatic mutations) and SVs with strong molecule support (remove false positives due to assembly errors). Somatic variability was investigated in the de novo assemblies by aligning single-molecule data to each map containing the repeat expansion. These molecules were manually visualized and subsequently plotted by bp distance between the two labels of interest that encompass the repeat expansion to show the presence/absence of somatic mosaicism (**Figure 3, Supplemental Table 3**).

### In situ hybridization with RNAScope

Human skin fibroblasts were derived from 4-mm skin biopsies, maintained, and banked as we previously described.^17^ For the fibroblast cell culture, 8-well chamber slides (Nunc Lab-Tek chamber slide, Sigma, Cat. No. C7182) were coated with 0.1% gelatin for 15 minutes. Sixty thousand cells were seeded in each chamber and incubated overnight. The next day, slides were fixed with 10% neutral buffered formalin for 15 minutes and dehydrated by gradually increasing ethanol concentrations (50%, 70%, 100%).

In situ hybridization was performed according to the manufacturer’s instructions using the RNAscope multiplex fluorescent assay v.2 kit. In brief, slides were treated with RNAScope hydrogen peroxide (Part Number 322335)) for 10 min at room temperature, followed by a protease III incubation (10 min, room temperature). Probe hybridization (Hs-*ATXN10*-ATTCT-C1, ACDBio, part ID 1001381), positive control (part number, 320861), and negative control (ACD, part number 320871) was performed in a HybEZ oven (ACD, HybEZ™ II Hybridization System, part number 321710) at 40°C for 2 hours. Slides were washed and underwent three steps of amplification: AMP1 (30 min, 40°C), AMP2 (40°C), and AMP3 (15 min, 40°C). Then, slides were incubated with RNAScope HRP-C1 for 15 minutes at 40°C, followed by a washing cycle and incubation with TSA Plus fluorophore (1:600) for 30 minutes at 40°C. Lastly, slides were blocked with a horseradish peroxidase (HRP) blocker for 15 minutes at 40°C and counterstained with a nuclear marker (DAPI). RNAScope slides were covered with Prolong Gold Antifade reagent (Invitrogen, P36930) and a 22 × 50 mm coverslip for imaging. Imaging was performed on ImageXpress Pico (Molecular Devices) and analyzed with CellReporterXpress Image acquisition software (Molecular devices version 2.6.130), and RNA foci were analyzed using the “pits and vesicles” software. Statistical analysis of RNA foci counts was performed in GraphPad Prism using a one-way ANOVA (α≤0.05) and a Brown-Forsythe & Welch test (95% CI).

## Results

We describe a four-generation Mexican family with an autosomal-dominant inheritance of an *ATXN10* repeat expansion. We recently reported on this family and, interestingly, identified an individual of this family affected with typical L-Dopa responsive Parkinson’s disease and a pure (ATTCT)_n_ repeat expansion^15^ (**Figure 1**). Here, we expanded our genetic studies and included additional techniques of optical genome mapping and RNAScope *in situ* hybridization to understand the repeat size, genomic structure, and somatic variability in order to gain more insight into the genetic mechanisms of the *ATXN10* repeat expansion, clinical presentation, and disease trajectory.

**Figure 1.**
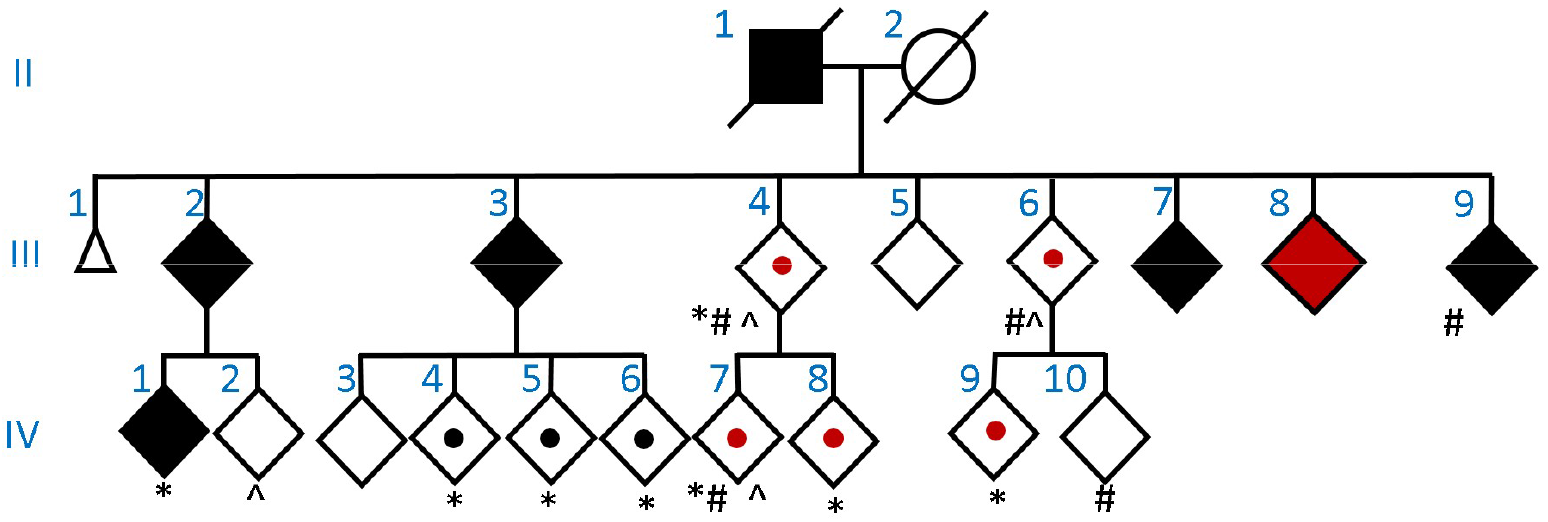
*ATXN10* Pedigree of Mexican family. Roman numerals indicate generations to the left of each pedigree. Diamond represent male or female, triangle indicates miscarriage, black symbols indicate affected individuals with SCA10, the red symbol indicates Parkinson’s disease, dots in the middle of the symbol mean *ATXN10* repeat expansion carrier, but unaffected. Black dots indicate carrier for interrupted repeat expansion, and the red dot indicates pure ATTCT repeat expansion. * indicates Cas9/SMRT sequencing, # indicates Saphyr optical genome mapping. The age range in generation III is 55-75 years, and in generation IV is 15-55 years. Extended pedigree with generations I and II is shown in Schüle et al. 2017^15^.

### Pure (ATTCT)_n_ repeat expansion carriers show reduced clinical penetrance for SCA10

We examined 22 family members from generations III (7 individuals) and IV (15 individuals) of the family (**Figure 1**) and collected blood and skin biopsies from individuals who consented to the study protocol. Sixteen individuals had an *ATXN10* repeat expansion (rs60726084, seven in generation III, nine in generation IV). We found six non-carriers in generation IV. Of the 16 carriers, ten presented with the mixed (ATTCT)_n_-(ATTCC)_n_ repeat expansion (black symbols, **Figure 1**), and six carried the pure (ATTCT)_n_ repeat expansion (red symbols, **Figure 1**). Of the ten cases with a mixed repeat expansion, five were affected with typical SCA10 with age at onset of 41.6 years (range 35-48 years, III.2, III.3, III.7, III.9, IV.1), and three of the five SCA10 cases had seizures (III.2, III.3, III.7). The five unaffected carriers of the mixed repeat could be pre-symptomatic are were between 15-20 and 51-55 at the time of examination. Further longitudinal clinical follow-up will be necessary.

Of the six relatives with pure (ATTCT)_n_ expansion, one developed Parkinson’s disease in late 30’s, while the other five were unaffected. Two of them have passed the average age at onset in this family by over 20 years (age a last visit between 65-70 years), while the other three carriers of the pure (ATTCT)_n_ repeat expansion could still be pre-symptomatic at age 15-20 and 40-44 years. None of the individuals with the pure (ATTCT)_n_ expansion presented with medical history or symptoms compatible with the diagnosis of SCA10 (**Supplemental Materials, case history III.6**).

### Inheritance of pure and mixed *ATXN10* repeat expansion

We observed two structurally very different repeat expansions in this family. One *ATXN10* repeat presented as a pure (ATTCT)_n_ expansion and the other as a mixed (ATTCT)_n_(ATTCC)_n_ expansion. There is no clear explanation of how a repeat can mutate from one complex repeat pattern to another in one generation, e.g., from generation II to III. It is also unusual that all siblings in generation III carry an extended *ATXN10* allele. Usually, in an autosomal-dominant inheritance pattern, one would expect 50% *ATXN10* repeat expansion carriers and 50% carriers of the normal allele. To exclude non-paternity, we tested 15 microsatellite markers, and all siblings in generation III tested showed matching alleles (**Supplemental Table 1**). Furthermore, we did not detect an expanded allele in the mother II.2 with No-Amp targeted sequencing. DNA from the father (II.1), who died before the study began, was unavailable.

Our interpretation from these clinical genetic findings is that the father was compound heterozygous for both the pure and the mixed extended allele. Clinically, the father presented with a typical SCA10 symptom complex from the medical history, including seizures and no aggravated clinical phenotype or faster progression. He died in his late 60’s.

### Accurate sizing of *ATXN10* pentanucleotide expansions

Previously, we used No-Amp targeted sequencing to sequence the entire *ATXN10* repeat expansion and detected expansions between 1076 and 1363 pentanucleotide repeat expansions (5,380bp to 6,815bp) in generation III of this family^15^. Long read No-Amp targeted sequencing allows for accurate sizing in repeat composition by sequencing through the entire *ATXN10* repeat without prior DNA template amplification. While we had confidence that the repeat length was correct, as the Cas9/small guide RNA recognition site is upstream of the repeat expansion, it was in stark contrast to the diagnostic sizing with PCR/Southern blotting by Athena Diagnostics. The proband III.8 with Parkinson’s disease presented 1304 repeats (6,520bp) for the expanded allele with PacBio. In contrast, diagnostic testing using Athena Diagnostics (Test code 6901, PCR, Southern Blot) resulted in 1968 ATTCT repeats (9,840bp) for the extended allele, a size difference of 664 repeats (3,320bp). Similarly, the affected brother III.7 presented 1363 repeats (6,815bp) with PacBio and 2223 ATTCT repeats of the extended allele (11,115bp) with Athena Diagnostics, a size difference of 860 repeats (4,300bp).

It is known that Southern blotting can result in variable sizing between different diagnostic labs; hence, we wanted to address the repeat sizing with another independent technique using an amplification-free strategy. We decided to perform optical genome mapping using high-density nanochannel arrays and imaged on the Saphyr™ instrument (Bionano Genomics) that allows the detection of structural variants greater than 500bp. Optical genome mapping images of long amplification-free DNA molecules fluorescently labeled at specific 6-7 bp sites preserve all genomic structural information. Optical genome mapping detects structural variants down to 1% variant allele fraction for mosaic samples. We collected new blood samples from four family members with known repeat expansions previously seen with No-Amp targeted sequencing.

All four samples with an extended allele were correctly categorized as an insertion in intron 9 of the *ATXN10* gene with coordinates chr22:46191235-46191304 (Hg38). Per sample, we resolved between 66 and 144 molecules (expanded alleles between 33 and 54) (**Supplemental Table 2**). The repeat expansion sizes were comparable between the NoAmp sequencing and optical genome mapping and showed similar sizing and variability.

### Somatic mosaicism was detected with both No-Amp targeted sequencing and optical genome mapping

Next, we compared the size range of the expanded allele for No-Amp targeted sequencing and optical genome mapping. For No-Amp targeted sequencing, we sequenced on average 101 CCS reads for the expanded allele (range 5 to 419 CCS reads). Interestingly, we found that samples with a mixed repeat show less variability in the size of the expanded allele (average of 1,539bp; range 177bp to 3,074bp) compared to the pure repeat expansion (average of 6,363bp; range 2,437 to 13,708bp). These data indicate that the pure expansion might be less stable and exhibit more somatic variability than the mixed *ATXN10* repeat expansion.

For Bionano optical genome mapping, we had two samples with a pure repeat expansion for direct comparison. While optical genome mapping cannot give precise estimates down to the bp resolution, we estimate from the consensus map size and subtract the size (5,355bp) between the consensus map labels to report the repeat expansion size. Both samples III.4 and IV.7 showed very similar repeat expansions and levels of variability (**Figures 2, 3, and 4N**). Interestingly, IV.7 showed a high degree of somatic mosaicism, which we further evaluated in skin fibroblasts from these individuals (**Figure 4**).

**Figure 2.**
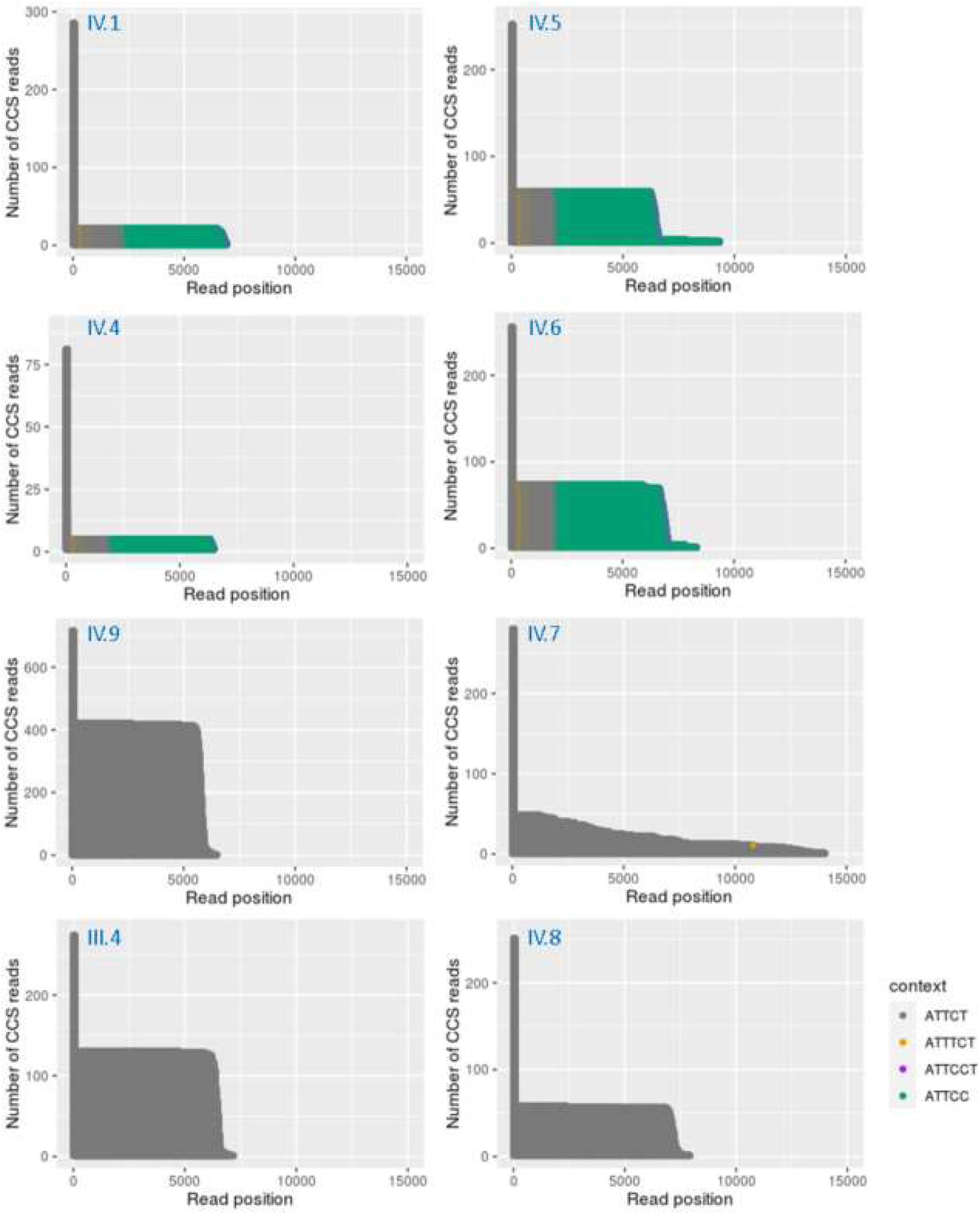
Waterfall plots for No-Amp targeted sequencing of ATXN10 carriers with pure (ATTCT)_n_ expansions and mixed (ATTCT)_n_-(ATTCC)_n_ expansions. A: Examples of pure and mixed repeat expansion from different family members in generations III and IV show inter and intra-individual variability. A row of dots represents repeat sequences in each sequencing read in this plot. Dots with different colors represent a different unit of repeat sequence context. Dark gray: ATTCT, Gold: ATTCC, Light blue: ATTTCT, Green: ATTCCT. B: Violin plots for ATXN10 repeat lengths distribution detected with No-Amp targeted sequencing.

**Figure 3.**
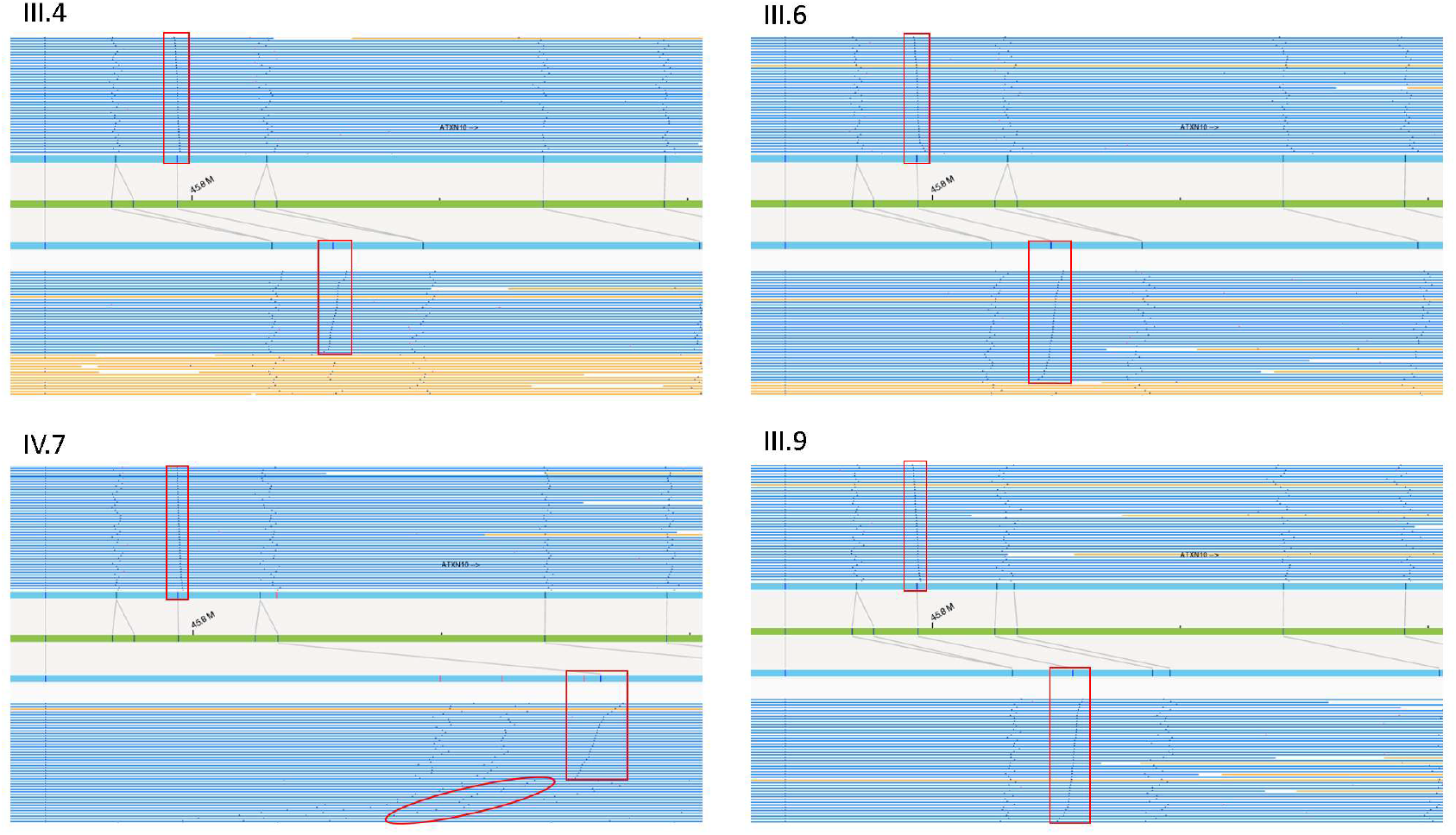
Visualization of optical genome mapping for four ATXN10 carriers as molecule distance graphs. The upper half of individual panels represents unexpanded molecules, and the lower part shows expanded alleles. The green bar represents the reference allele (hg38) with marks at all locations of the sequence motif (vertical lines), which is recognized by the Bionano optical genome mapping enzyme (e.g., DLE-1, Nt.BspQI, Nb.BssSI), which occurs every five kbp on average. Genome maps (blue bars) are produced from images of labeled DNA molecules >150 kbp for the target DNA. **We measured expanded alleles between reference label IDs 5**,**045 and 5**,**048 (Chromosome 22, Hg38)**.

**Figure 4.**
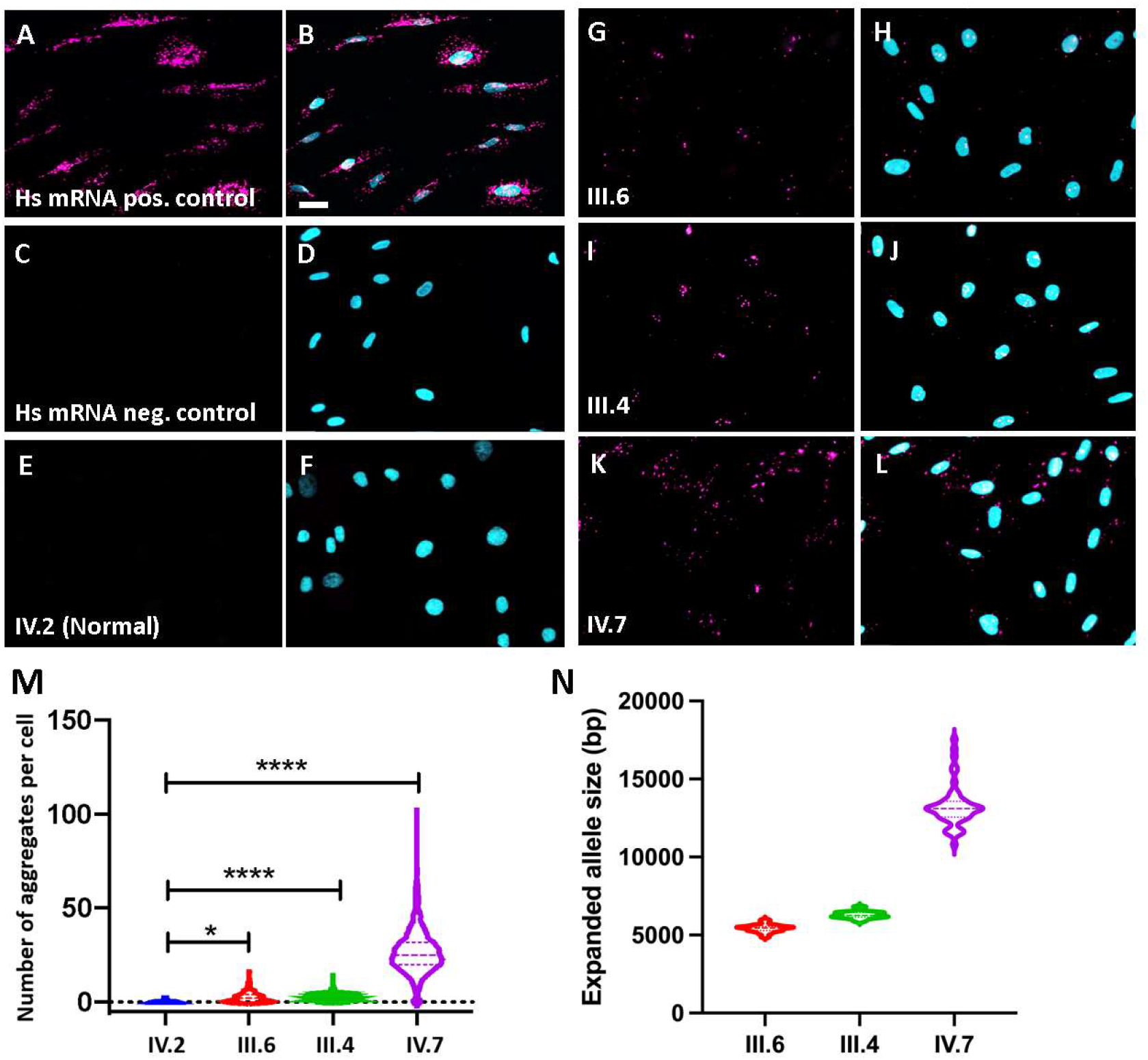
Figure. A-L: RNAScope in situ hybridization of *ATXN10* repeat expansion. A-B: RNAscope human mRNA positive control. C-D: RNAscope human negative control. E-F: Healthy control, G-H: *ATXN10* expansion carrier III.6, I-J: *ATXN10* expansion carrier III.4, K-L: *ATXN10* expansion carrier IV.7. E, G, I, K: Hs-*ATXN10*-ATTCT-C1 RNA Scope probe; B, D, F, H, J, L: Merge with DAPI. The scale bar in B represents 20 um. **M: Violin plots of counts of RNA foci per cell using ImageXpress**. Mean number of RNA signals per cells for control =0.29, SD 0.6 (1870 nuclei), III.6 =3.51, SD 2.33 (2034 nuclei), III.4 =3.89, SD 2.70 (759 nuclei), IV.7 =26.65, SD 11.71 (2034 nuclei). **N: Violin plots for Bionano Saphyr amplification-free optical genome mapping structural variant calls** of the *ATXN10* repeat expansion region for the same *ATXN10* repeat expansion carriers (bp = base pairs). Median expanded allele size III.6 =5,459bp (≜1,091 ATTCT repeats), III.4 =6,260bp (≜1,252 ATTCT repeats), IV.7 =13,102bp (≜ 2620 ATTCT repeats). P-values are represented as * with *=0.01, ****=0.0001 Deciphering structural composition of *ATXN10* repeat expansion

### Variable *ATXN10* repeat sizes correlate with varying counts of *ATXN10* RNA foci in patient-derived skin fibroblasts

To investigate to what extent the somatic variability of the *ATXN10* repeat size relates to (ATTCT)_n_ repeat RNA foci within cells, we custom-designed an in situ hybridization RNAScope® probe against the ATTCT repeat. Spliced intronic repeat expansions can accumulate as intracellular RNA foci, contributing to cellular stress and neurodegeneration.

We derived skin fibroblasts from three *ATXN10* repeat expansion carriers (III.4, III.6, and IV.7) and one healthy relative (IV.2) with two normal *ATXN10* alleles. We detected RNA foci within nuclei and cytoplasm in all carriers with the expanded ATNxN10 allele and none in control (**Figure 4 E-L**). Positive and negative RNAScope controls were included (**Figure 4A-D**). When we quantified the number of RNA aggregates in the three carriers with the expanded *ATXN10* allele, we counted an average of 3.51 and 2.33 foci in cases III.4 and III.6 and 26.65 in case IV.7) with a wide range of counts that correlated to the No-Amp sequencing data and the *ATXN10* repeat sizing of the optical genome mapping (**Table 1, Figure 4M**).

**Table 1.** Allele count and sizes for normal and expanded *ATXN10* alleles in a Mexican family.

| Pedigree ID | Allele 1 count | Allele 1 size range (bp) | Allele 1 count range | Allele 2 count | Expanded Allele size range (bp) | Repeat type | Range of expanded allele size (bp) | ATTCT/ATTCC total count range | ATTCT count range | ATTCT count range difference | ATTCC count range | ATTCC count range difference |
| --- | --- | --- | --- | --- | --- | --- | --- | --- | --- | --- | --- | --- |
| IV.5 | 193 | 64-77 | 13-15 | 59 | 6244-9318 | imperfect | 3074 | 1172-1835 | 367-391 | 24 | 805-1444 | 639 |
| IV.6 | 183 | 64-73 | 13-15 | 73 | 5914-8314 | imperfect | 2400 | 1131-1640 | 362-398 | 36 | 769-1242 | 473 |
| IV.1 | 263 | 64-70 | 13-14 | 22 | 6422-6927 | imperfect | 505 | 1211-1363 | 407-443 | 36 | 804-920 | 116 |
| IV.4 | 76 | 75-80 | 15-16 | 5 | 6338-6515 | imperfect | 177 | 1235-1278 | 375-385 | 10 | 860-893 | 33 |
| III.4 | 143 | 64-70 | 13-14 | 130 | 4750-7187 | perfect | 2437 | 950-1435 | 950-1435 | 485 | none | none |
| IV.8 | 193 | 79-85 | 16-17 | 58 | 2383-7920 | perfect | 5537 | 476-1552 | 476-1552 | 1076 | none | none |
| IV.9 | 297 | 65-85 | 13-17 | 419 | 2730-6502 | perfect | 3772 | 546-1293 | 546-1291 | 745 | 0-2 | 2 |
| IV.7 | 271 | 74-83 | 15-17 | 49 | 330-14038 | perfect | 13708 | 66-2782 | 66-2782 | 2716 | none | none |

## Discussion

Three factors of the repeat expansion might influence clinical presentation and outcome: repeat length, repeat interruption, and somatic mosaicism^18; 19^.

While it is known that the pathogenic repeat length for the ATXN10 repeat expansion is >850 repeats, there are significant gaps in our current knowledge about the nature and clinical relevance of repeat composition and repeat interruptions. Advanced long-read sequencing technologies and optical genome mapping allow us now to fully resolve very large repeat expansions.

### Somatic mosaicism of (ATTCT)_n_ and (ATTCT)_n_(ATTCC)_n_ repeat expansions

The advantage of No-Amp targeted sequencing, and optical genome mapping is that these techniques directly use genomic DNA without introducing amplification errors, recombination effects, or cell culture artifacts through prior amplification.

On the one hand, we found those repeat interruptions, as in the mixed (ATTCT)n(ATTCC)n repeat expansions, can stabilize the repeat as been previously reported (Figure 2)^24^. On the other hand, the carriers with the pure (ATTCT)_n_ expansion show more variability in the size of the expanded alleles (average 1255 (range 48 to 2716 ATTCT repeats) compared to the mixed repeat expansion alleles (average 315 (range 33 to 639 ATTCT repeats). In particular, we identified one carrier (IV.7) with a very unstable (ATTCT)_n_ allele ranging from 66-2782 (ATTCT)_n_ repeats. This variability is present with NoAmp Sequencing and Bionano optical genome mapping (**Figure 2, 3**).

This unstable repeat expansion also translates into variable counts of intracellular inclusions or RNA foci in primary skin fibroblasts from these cases. However, we do not know if and how these RNA foci translate to cellular stress, degeneration, and the development of clinical symptoms. Still, it has been shown that specific RNA binding proteins get sequestered by the repeat expansion with particular sequence motifs, e.g., heterogeneous nuclear ribonucleoprotein K binds to the ATTCT repeat^25^.

### Reduced disease penetrance in pure ATTCT carriers

In this Mexican kindred, we identified two distinct *ATXN10* repeat expansions. One expansion was a pure ATTCT_(n)_ expansion with no interruptions and a repeat length between 1278 and 2782 repeats (**Supplemental Figure 1**). The second repeat is a mixed repeat with ATTCT_(n)_ ATTCC_(n)_ clinically associated with SCA10, whereas the pure repeat expansion shows reduced penetrance for SCA10.

Of the six individuals with pure *ATXN10* repeat expansions, none showed clinical signs that would allow for the diagnosis of SCA10. However, as we reported earlier, one relative presented with Parkinson’s disease^15^. While three individuals with expansions might be pre-symptomatic (at ages 15-20 and 40-44 years), two relatives from the third generation were between 65 and 70 years at the time of assessment. They did not show clinical signs of SCA10 (assessed by F.J.J.G., A.C-P., clinical examination III.6 in **Supplemental Material**).

Notably, all siblings tested (7 of 8 siblings) in generation III carry either a pure or a mixed repeat expansion, suggesting that their father II.1 was compound heterozygous for the pure and mixed *ATXN10* repeat expansion. By history, he had the typical presentation of SCA10 and died in his late 60’s. The combination of both repeats did not seem to present with a more severe clinical phenotype.

Both results suggest that the pure originally described ATTCT_(n)_ expansion is not pathogenic, whereas repeat insertions or interruptions cause SCA10 symptoms. Only since it is possible to sequence through the entire repeat expansion from genomic DNA with long-read sequencing techniques is it possible to resolve the complete sequence of the ATXN10 repeat expansion fully, as it is not possible to decipher the sequence composition using Southern blot analysis, PCR electrophoresis or repeat-prime PCR^13^.

Therefore, *ATXN10* repeat expansions fall into a category of repeat expansions that only become clinically symptomatic when repeat interruptions are present. Similar genetic loci have recently been reported for SCA31 (*BEAN1*)^20^, SCA37 (*DAB1)*^21; 22^, and three loci for benign adult familial myoclonic epilepsy BAFME (SAMD12, TNRC6A, RAPGEF2)^23^, where repeat interruptions are inserted in non-pathogenic repetitive motifs.

In SCA31, that is caused by a complex intronic pentanucleotide repeat-containing (TGGAA)n, (TAGAA)_n_, (TAAAA)_n_, and (TAAAATAGAA)_n_ the BEAN1 (brain expressed, associated with Nedd4) gene, only the (TGGAA)_n_ repeat drives pathogenicity^20^. An (ATTTC)_n_ insertion causes SCA37 within a polymorphic ATTTT repeat in the non-coding in the 5’-untranslated region of the *DAB1* gene (Disabled-1 gene)^21; 22^. Lastly, benign adult familial myoclonic epilepsy (BAFME) is caused by intronic (TTTCA)_n_ and (TTTTA) _n_ repeats in the SAMD12 (sterile α-motif domain–containing 12), TNRC6A (trinucleotide repeat-containing 6A) and RAPGEF2 (Rap guanine nucleotide– exchange factor 2) genes, where the same repeat (TTTCA)_n_ and (TTTTA) _n_ is driving pathogenicity regardless of the locus or gene in which the repeats are located^23^.

We propose that the pure ATTCT_(n)_ is non-pathogenic or has only low penetrance with a mild disease course. In contrast, repeat interruptions mainly drive the clinical symptoms and presumably neuropathology. Longitudinal follow-up and *in vitro* functional studies will be essential to understand the ATXN10 repeat expansion pathophysiology further. Mechanistically, the insertion of repeats into the (ATTCT)_n_ might be a driving factor for disease causation in SCA10.

In summary, No-Amp targeted sequencing technology and optical genome mapping allow for accurate sizing of the repeat expansion and composition and reveal somatic mosaicism, which is impossible with current diagnostic methods. Deciphering the composition of repeat expansion will allow us to distinguish between pathogenic and non-pathogenic repeats, unravel disease mechanisms, and develop targeted therapies. These advanced genetic technologies are critical in implementing diagnostic testing to diagnose disease better, and accurate genetic counseling as variation in the repeat can affect disease penetrance, symptoms, and disease trajectory.

## Data Availability

All data produced in the present study are available upon reasonable request to the authors.

## Appendices

### Supplemental Information description

Supplemental data include case history, supplemental tables, and figures.

## Acknowledgments

We are indebted to the family members for their participation in this study and their commitment to helping the understanding of underlying genetic causes of neurodegenerative disorders. We thank Jose Miranda for supporting study recruitment. Start-up funding and Chan Zuckerberg Initiative Supplement funds to B.S. supported the research. We received in-kind donations from PacBio and Bionano in the form of sequencing and optical genome mapping.

## Declaration of Interests

I. McLaughlin, Y.-C. Tsai, J. S. Ziegle are employees and shareholders of Pacific Biosciences. K. Hong, J. C. Y. Lai, J. Lee, and M. Gallagher are employees and shareholders of Bionano Genomics. B. Schüle, F. Jimenez Gil, J. Palafox Vasquez, F. Zafar, C. A. Morato Torres, F. Torres, T. Ashizawa, J. Fernandez-Ruiz, A. Chirino-Perez, A.O. Romero-Molina declare no competing interests.

## Notes

### Funding Statement

Chan Zuckerberg Initiative supplemental funding

