## Supplemental Material for "ATTCT and ATTCC repeat expansions in the *ATXN10* gene affect disease penetrance of spinocerebellar ataxia type 10"

[1]Department Pathology, Stanford University School of Medicine, Stanford, CA 94305, USA
[2]Pacific Biosciences of California, Inc, Menlo Park, CA, 94025, USA
[3]Bionano Genomics, San Diego, CA, 92121, USA
[4] Laboratorio de Neuropsicología, Departamento de Fisiología, Facultad de Medicina, Universidad Nacional Autónoma de México, CDMX, Mexico
[5]Southern California Permanente Medical Group, Oxnard, California, 93036, USA
[6]Department Neurology, Houston Methodist Research Institute, Houston, Texas, 77030, USA
[7]Hospital San Javier S.A. de C.V., Guadalajara, Jalisco, 44670, Mexico
¶These authors contributed equally to this work.

**Case History for III.6 available upon reasonable request.**

Supplemental figures and table.

**Supplemental Table 1. AmpFlSTR™ Identifiler™ testing**

| Pedigree IDs | FAM | | | | | | | | VIC | | | | | | | | | |
|---|---|---|---|---|---|---|---|---|---|---|---|---|---|---|---|---|---|---|
| | D8S1179 | | D21S11 | | D7S820 | | CSF1PO | | D3S1358 | | TH01 | | D13S317 | | D16S539 | | D2S1338 | |
| II.1 (inferred) | 151 | 142 | 214 | 221 | 275 | | 326 | 330 | 129 | | 187 | 176 | 225 | | 272 | 279 | 329 | 337 |
| II.2 | 142 | 159 | 206 | 221 | 271 | 275 | 330 | | 129 | 143 | 176 | 180 | 229 | 238 | 279 | 279 | 316 | 316 |
| III.2 | 142 | 151 | 206 | 214 | 275 | 275 | 330 | 330 | 129 | 143 | 180 | 187 | | | 272 | 279 | 316 | 337 |
| III.4 | 142 | 142 | 221 | 221 | 271 | 275 | 326 | 330 | 129 | 129 | 176 | 187 | 225 | 238 | 272 | 279 | 316 | 337 |
| III.6 | 151 | 159 | 206 | 221 | 275 | 275 | 330 | 330 | 129 | 143 | 180 | 187 | | | 272 | 279 | 316 | 337 |
| III.7 | 142 | 159 | 206 | 214 | 271 | 275 | 330 | 330 | 129 | 143 | 180 | 187 | 225 | 229 | 279 | 279 | 316 | 329 |
| III.8 | 151 | 159 | 214 | 220.9 | 275 | 275 | 330 | 330 | 129 | 143 | 176 | 180 | 225 | 229 | 272 | 279 | 316 | 329 |
| III.9 | 142 | 159 | 206 | 221 | 271 | 275 | 330 | 330 | 129 | 129 | 176 | 176 | 225 | 229 | 272 | 279 | 316 | 337 |

| Pedigree IDs | NED | | | | | | | | PET | | | | | |
|---|---|---|---|---|---|---|---|---|---|---|---|---|---|---|
| | D19S33 | | vWA | | TPOX | | D18S51 | | Amelogenin (X/Y) | | D5S818 | | FGA | |
| II.1 (inferred) | 123 | 121 | 180 | 188 | 234 | 242 | 306 | 298 | | 113 | 139 | 153 | 241 | 245 |
| II.2 | 121 | 129 | 176 | 180 | 234 | 234 | 286 | 294 | 108 | | 153 | 153 | 237 | 254 |
| III.2 | 121 | 123 | 176 | 180 | 234 | 234 | 286 | 298 | 108 | | 139 | 153 | 241 | 254 |
| III.4 | 121 | 121 | 176 | 188 | 234 | 234 | 294 | 298 | 108 | 113 | 139 | 153 | 237 | 245 |
| III.6 | 123 | 129 | 176 | 188 | 234 | 234 | 294 | 298 | 108 | | 139 | 153 | 241 | 254 |
| III.7 | 121 | 123 | 176 | 188 | 234 | 234 | 286 | 298 | 108 | 113 | 139 | 153 | 245 | 254 |
| III.8 | 121 | 121 | 176 | 188 | 234 | 242 | 294 | 306 | 108 | 113 | 139 | 153 | 237 | 245 |
| III.9 | 121 | 129 | 180 | 188 | 234 | 242 | 286 | 298 | 108 | | 153 | 153 | 237 | 241 |

**Supplemental Table 2. Summary statistics and QC for optical mapping for de novo assembly pipeline.**

| **Sample** | **III.9** | **III.6** | **IV.10** | **III.4** | **IV.7** |
|---|---|---|---|---|---|
| Data collected (molecules >150kbp) | 1,314.1 Gbp | 1,327.86 Gbp | 1,336.59 Gbp | 1,325.12 Gbp | 1,348.2 Gbp |
| Molecule N50 | 279.75 kbp | 300.38 kbp | 269.25 kbp | 251.25 kbp | 308.63 kbp |
| Diploid genome map length | 6,036.48 Mbp | 6,067.1 Mbp | 6,003.15 Mbp | 6,095.06 Mbp | 6,009.44 Mbp |
| Diploid genome map N50 | 80.4 kbp | 59.72 kbp | 69.69 kbp | 70.44 kbp | 73.6 kbp |
| Total ATXN10 allele counts | 66 | 132 | 122 | 111 | 144 |
| Expanded allele counts | 33 | 54 | 0 | 51 | 48 |
| ***de novo assembly pipeline SV calls against hg38*** | | | | | |
| Insertions | 2813 | 2853 | 2876 | 2862 | 2904 |
| Deletions | 1313 | 1286 | 1275 | 1269 | 1255 |
| Inversion breakpoints | 53 | 59 | 49 | 64 | 58 |
| Duplications | 76 | 82 | 78 | 86 | 72 |
| Translocations | 7 | 9 | 10 | 6 | 8 |

**Supplemental Figure 1.**

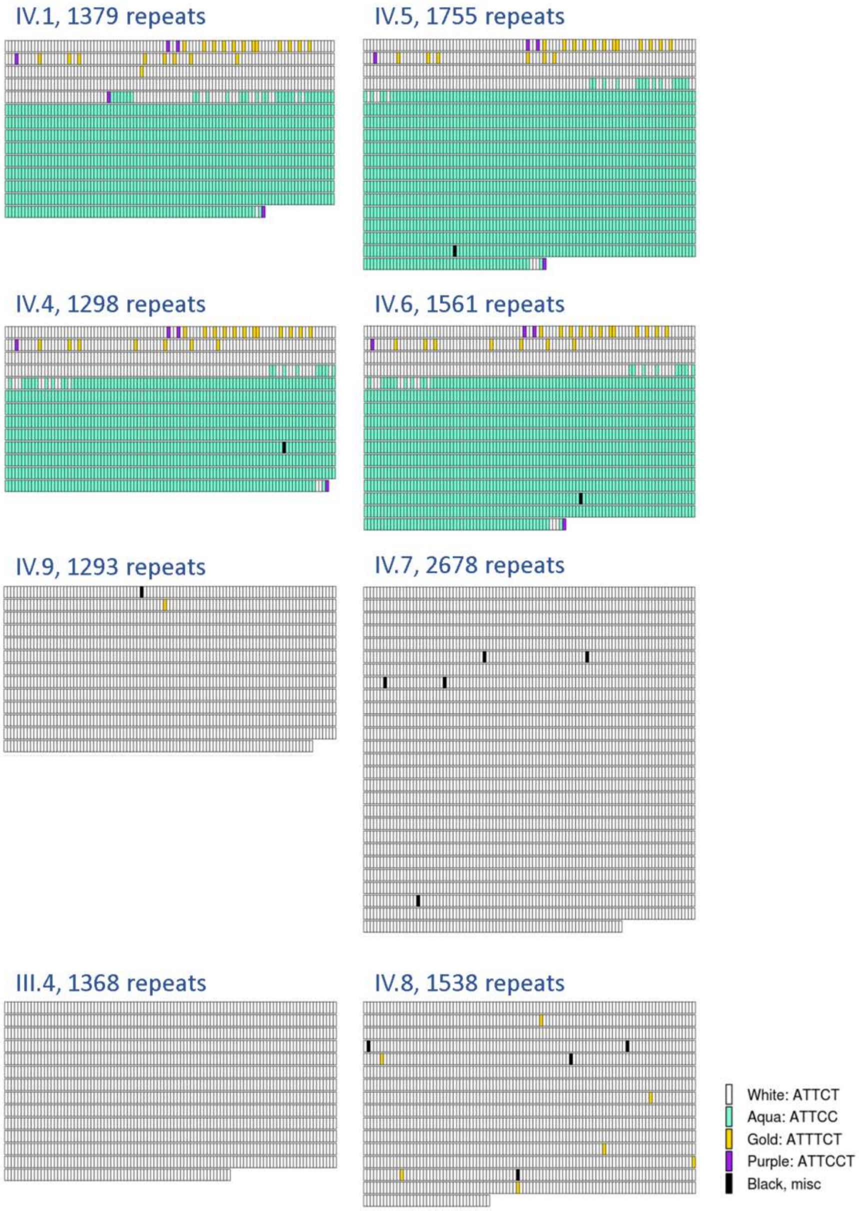


**Supplemental Figure 1. *ATXN10* repeat expansion sequence schematics based on No Amp sequencing.** Repeat expansions are represented in the 5′ (upper left) to 3′ (lower right) direction. The schematics and expansion size given are based on the most error-free read length for each sample. Each rectangle represents a sequence motif as follows: white, ATTCT; aqua, ATTCC; gold, ATTTCT; purple, ATTCCT. Black rectangles denote miscellaneous motifs that may represent errors in sequence reads.
